# Sustained improvements in brain health and metabolic markers 24 months following bariatric surgery

**DOI:** 10.1101/2023.03.08.23286944

**Authors:** Marianne Legault, Mélissa Pelletier, Amélie Lachance, Marie-Ève Lachance, Yashar Zeighami, Marie-Frédérique Gauthier, Sylvain Iceta, Laurent Biertho, Stephanie Fulton, Denis Richard, Alain Dagher, André Tchernof, Mahsa Dadar, Andréanne Michaud

**Author notes:** co-last authors (Mahsa Dadar, Andréanne Michaud).

## Abstract

**Background:** Obesity and its metabolic complications are associated with lower gray matter (GM) and white matter (WM) density, whereas weight loss after bariatric surgery leads to an increase in both measures. These increases of GM and WM density are significantly associated with post-operative weight loss and improvement of the metabolic/inflammatory profiles. While our recent studies demonstrated widespread increases in WM density 4 and 12 months after bariatric surgery, it is not clear if theses changes persist over time. The underlying mechanisms also remain unknown. In this regard, numerous studies demonstrate that the enlargement or hypertrophy of mature adipocytes, particularly in the visceral fat compartment, is an important marker of adipose tissue dysfunction and obesity-related cardiometabolic abnormalities.

**Objective:** To assess whether the previously observed increases in WM and GM densities are maintained 24 months post-surgery, and to examine if pre-operative abdominal omental (OM) and subcutaneous (SC) adipocyte diameters are associated with WM and GM changes after bariatric surgery.

**Methods:** 32 participants undergoing bariatric surgery were recruited. WM and GM densities were assessed from T1-weighted MRIs acquired prior to and 4-, 12-, and 24-months post-surgery using voxel-based morphometry. OM and SC adipose tissue samples were collected during the surgical procedure. OM and SC adipocyte diameters were measured by microscopy of fixed adipose tissue samples. Linear mixed-effects models were performed controlling for age, sex, surgery type, initial BMI, and diabetic status.

**Results:** The average weight loss at 24 months was 33.5±7.2%. A widespread increase in WM density was observed 24 months post-surgery mainly in the cerebellum, brainstem, and corpus callosum (p<0.05, FDR) as well as some regions in GM density. Greater baseline OM adipocyte diameter was associated with greater changes in total WM density at 24 months (p=0.008). A positive trend was observed between SC adipocyte diameter and changes in total WM density at 24 months (p=0.05).

**Conclusion:** Our results show prolonged increases of WM and GM densities up to 24 months post-bariatric surgery. Greater preoperative OM adipocyte diameter is associated with greater increases in WM density at 24 months, suggesting that individuals with excess visceral adiposity might benefit the most from surgery.

## 1. Introduction

Excessive accumulation of adipose tissue in the body, especially in the visceral fat compartment (mesentery and greater omentum), is strongly associated with cardiometabolic alterations, including insulin resistance, dyslipidemia, hypertension and chronic, low-grade inflammation [1], [2]. Recent studies highlighted that adipose tissue dysfunction, which leads to limited lipid storage and reduced adipose tissue expandability, is one of the major mechanisms explaining the link between visceral obesity and cardiometabolic alterations [3]. Numerous studies demonstrate that the enlargement or hypertrophy of mature adipocytes, particularly in the visceral fat compartment, is an important marker of adipose tissue dysfunction [4] and obesity-related cardiometabolic abnormalities [5].

In addition to these cardiometabolic alterations, overall and abdominal obesity in midlife are risk factors for cognitive impairment and dementia [6]–[8]. Previous cross-sectional and longitudinal magnetic resonance imaging (MRI) studies demonstrated that obesity is associated with reduced grey matter (GM) volume and lower cortical thickness, mainly in fronto-temporal regions [9]–[13]. Other studies have also shown that adiposity is related to increased white matter hyperintensities, which were in turn relate to lower cortical thickness and diminished GM volume [14], [15]. A meta-analysis by Daoust et al. [16] highlighted that obesity is associated with reduced white matter (WM) integrity in a tract linking frontal areas involved in executive function. These alterations in brain integrity might explain the association between abdominal obesity, accelerating brain aging and cognitive impairment. Even if the exact mechanisms underlying these obesity-related brain abnormalities are poorly characterized, there is an increasing number of studies showing that abdominal obesity-related cardiometabolic alterations, such as chronic, low-grade inflammation, hypertension, and insulin resistance, might have negative effects on cerebral GM and WM integrity by disrupting cerebral blood flow [17], leading to cerebral hypoperfusion and cognitive impairment [14], [17].

Bariatric surgery is an interesting model to examine the impact of marked weight loss and cardiometabolic improvements on brain health in a longitudinal setting [18]–[20]. The first results of our brain MRI study reported rapid and important structural and functional brain changes 4 and 12 months after bariatric surgery [21]. We observed widespread increases in WM and GM densities [21] as well as resting neural activity [22] 12 months after bariatric surgery, suggesting a global effect on brain health. Similar findings were also reported by other groups [23]–[26]. In parallel to the increase in GM density, we found an increase in spontaneous neural activity following surgery, as assessed by the fractional amplitude of low frequency fluctuations (fALFF) signal [22]. These structural and functional increases were significantly associated with the magnitude of bariatric surgery-induced weight loss and metabolic improvements, suggesting that bariatric surgery may lead to a resolution of adiposity-related brain abnormalities along with widespread improvements in metabolism [21], [22]. Despite the growing evidence showing seemingly beneficial brain structural changes after bariatric surgery, it is not clear if these changes persist longer than 12 months [21]–[24]. In addition, the mechanisms explaining these changes are still unknown.

The objectives of the present study were i) to assess whether the increases in WM and GM densities previously observed at 12 months are maintained 24 months after surgery, ii) to examine the association between these structural brain changes and adiposity and metabolic markers 24 months after bariatric surgery, and iii) to examine the association between abdominal adipocyte diameter at the time of surgery and post-surgery GM and WM density changes. Finally, to verify that the changes in WM and GM densities observed after bariatric surgery are not due to any confounders related to the repetition of the MRI session and are related to the weight-loss induced-bariatric surgery, a separate group of participants matched with the surgery group for initial age and BMI underwent two scanning sessions before the surgery and one after.

## 2. Methods

### 2.1 Participant recruitment

Patients with severe obesity (n=32, 24 women, 8 men, mean age = 46.9 ± 7.6 years, mean BMI = 44.0 ± 4.3 kg/m²) scheduled to receive bariatric surgery at *Institut universitaire de cardiologie et pneumologie de Québec - Université Laval* (IUCPQ-UL) were recruited. Participants were tested before and 4, 12 and 24 months after surgery. Inclusion criteria were the following: 1) women or men with a BMI ≥35 kg/m² who require intestinal surgery, and who meet the NIH Guidelines for bariatric surgery [27]; 2) age between 18 and 60 years. Exclusion criteria were the following: 1) BMI<35 kg/m²; 2) age under 18 or over 60 years; 3) any uncontrolled medical, surgical, neurological or psychiatric condition; 4) liver cirrhosis or albumin deficiency; 5) any medication that may affect the central nervous system; 6) pregnancy; 7) substance or alcohol abuse; 8) previous gastric, oesophageal, brain or bariatric surgery; 9) gastro-intestinal inflammatory disease or gastro-intestinal ulcers; 10) severe food allergy; and 11) contraindications to MRI (implanted medical device, metal fragment in body, or claustrophobia). The study was approved by the Research Ethics Committee of the *Centre de recherche de l’IUCPQ-UL*, and all participants provided written informed consent to participate in the study.

### 2.2 Surgical procedures

Participants underwent sleeve gastrectomy, Roux-en-Y gastric bypass or biliopancreatic diversion with duodenal switch. All surgeries were performed at the IUCPQ-UL. Most participants underwent sleeve gastrectomy (n=21), consisting of a 250 cm^3^ vertical gastrectomy starting 5 cm–7 cm from the pylorus to the Hiss angle, using a 34–44 French Bougie for guidance, to create a gastric tube [28]. The greater curvature and fundus of the stomach were removed. Seven participants underwent a laparoscopic Roux-en-Y gastric bypass in which a proximal gastric pouch of 30-50 cm^3^ is created and anastomosed to the proximal small bowel by bypassing the first 100 cm and bringing a 100 cm alimentary limb on the gastric pouch [26]. Four participants underwent biliopancreatic diversion with duodenal switch, which is a mixed-surgery combining restrictive and malabsorptive mechanisms by creating a 250 cm^3^ vertical sleeve gastrectomy and duodeno-ileal anastomosis 100 cm from ileocecal valve [29].

### 2.3 Study design and experimental procedures

The study design has been described in detail by Michaud et al. [21]. Briefly, participants were asked to attend 4 sessions: prior to and 4, 12 and 24 months after surgery. To make sure that the brain changes observed after bariatric surgery were not due to any confounders related to the repetition of the MRI session, we recruited a group of 19 participants (13 women, 6 men, mean age = 43.8 ± 10.2 years, mean BMI = 41.9 ± 3.8 kg/m²) who were tested two times prior to surgery, approximately 4 and 1 month before surgery. At each session, all the participants had a complete physical and medical evaluation (blood pressure, anthropometry i.e. weight by bioimpedance, height, waist, neck and hip circumference with standard procedures [21]) and a brain MRI session. BMI was calculated from weight and height and total weight loss was calculated by subtracting follow-up weight from baseline weight. Excess weight loss was calculated using preoperative weight, postoperative weight, and ideal body weight for a BMI of 23 kg/m^2^ as previously used [30]. Fasting blood samples were taken on the morning of each visit. We used EDTA-coated tubes or serum clot activator tubes. All samples were immediately placed at 4℃ and then centrifuged and stored at -80℃. The plasma lipid profile (cholesterol, high-density lipoproteins, low-density lipoproteins, and triglyceride levels) and glucose homeostasis (plasma insulin and glucose levels) were measured by the biochemistry department of the IUCPQ-UL. The homeostasis model assessment insulin resistance (HOMA-IR) index was calculated using this formula: (fasting glucose (mmol/L) x fasting insulin (pmol/L))/135.

### 2.4 T1-weighted MRI acquisition

MRI acquisition was conducted in the morning, and participants were asked to fast 12h before the scanning session. One hour before the MRI session, participants were given a standardized beverage meal (237 ml, Boost original, Nestle Health Science) to control for hunger level [21]. They were asked to drink it within a 5 min period. Hunger levels were rated before and after the MRI session using a visual analog scale. The MRI protocol included anatomical T1-weighted three-dimensional (3D) turbo field echo images, resting state fMRI and a task state fMRI for food-cue reactivity. In the current study, only results from anatomical images are presented. T1-weighted three-dimensional (3D) turbo field echo images were acquired at the initial visit, 4 months, 12 months, and 24 months after surgery using a 3T whole-body MRI scanner (Philips, Ingenia, Philips Medical Systems) equipped with a 32-channel head coil at the CRIUCPQ-UL. The following parameters were used: 176 sagittal 1.0 mm slices, repetition time/echo time (TR/TE) = 8.1/3.7 ms, field of view (FOV) = 240 × 240 mm^2^, and voxel size = 1 × 1 × 1 mm.

### 2.5 Voxel-based morphometry measurements

GM and WM densities were assessed from each T1-weighted MRI using a standard voxel-based morphometry (VBM) pipeline [10]. The preprocessing steps were the following: 1) image denoising [31]; 2) intensity non-uniformity correction [32]; and 3) image intensity normalization into range (0 to 100) using histogram matching. Images were then first linearly (using a nine-parameter rigid registration) and then nonlinearly registered to an average brain template (MNI ICBM152) as part of the ANIMAL software [33] and segmented into GM, WM and cerebrospinal fluid images. These steps remove global differences in the size and the shape of individual brains and transform individual WM or GM density maps to the standardized MNI ICBM152 template space. VBM analysis was performed using MNI MINC tools (http://www.bic.mni.mcgill.ca/ServicesSoftware/MINC) to generate WM and GM density maps representing the local WM/GM concentration per voxel. To avoid the overlap between GM and WM signal in the border of GM and WM (due to a combination of partial volume effect and smoothing of the maps), we removed three voxels on the border of the GM and WM regions.

### 2.6 Adipocyte diameter

Subcutaneous (SC) and OM adipose tissue samples (n=29) were collected during the surgical procedure at the site of incision (lower abdomen) and at the greater omentum, respectively. Adipose tissue samples were fixed as previously described [34]. Adipocyte size was measured using automated segmentation with imageJ (https://imagej.nih.gov/ij/download.html) based on the method of Maguire et al. [35]. Whole slides were first digitized using a Zeiss Axio Scan.Z1 in brithfield at 20X magnification. The images were then converted in black and white 8-bit images. The areas of each adipocyte cell were measured. The mean areas were converted in diameter in µm. Samples with under 200 cells counted (n=14) were manually validated to assess the quality of the tissue. Samples with extreme tissue damage (n= 3) were excluded.

### 2.7 Statistical analysis

Repeated-measures ANOVA or Student’s T-test were used to compare the clinical characteristics of participants at baseline, 4, 12, and 24 months after bariatric surgery. Linear mixed-effects models were used to assess VBM changes of WM and GM following bariatric surgery, with session as a categorical fixed variable allowing us to contrast differences across sessions (e.g. 24 months vs baseline) and subject as a categorical random variable. Age at baseline, sex, initial BMI, baseline diabetic status, and surgery type were included in the model as covariates, while the session was a fixed effect. The mixed-effects model estimates are represented by t statistics in the results section. The VBM results were corrected for multiple comparisons using the False Discovery Rate (FDR) controlling technique with a significance threshold of p<0.05. To examine the associations with adiposity and metabolic variables and surgery-induced brain changes in GM or WM density, the AAL Atlas [36] was used to extract average regional VBM GM values and similarly, the Atlas from Yeh et al. [37] was used to extract regional VBM WM values for each participant. Mean GM and WM densities were extracted across all the participants. Linear mixed-effects models were then used to assess the association between post-surgery changes in GM or WM density and changes in adiposity and metabolic variables. We also tested the model including diabetic status, baseline BMI, and surgery type as covariates in the models to ensure the stability of the findings. Linear mixed-effects models were also performed to examine the association between OM or SC adipocyte diameter at the time of surgery and changes in total GM or WM density. Age and sex were included in the linear mixed-effects models as covariates and subject ID was included as a categorical random effect. Spearman correlations were performed to examine the associations between OM and SC adipocyte diameters and metabolic/adiposity variables. Bonferroni correction was applied for multiple comparisons. To control for re-test effect, paired t tests were used to compare data between the two sessions prior to surgery. Statistical analyses were performed with JMP software version 14 (SAS Institute Inc, Cary, NC, USA) and MATLAB software version R2021a (Natick, Massachusetts: The MathWorks Inc.)

## 3 Results

### 3.1 Clinical characteristics of participants

Clinical characteristics of participants are shown in **Table 1**. Participants were mainly women (75%) and the mean age at baseline was 46.9 ± 7.6 years. Nine participants suffered from type 2 diabetes prior to surgery. As expected, BMI, waist and neck circumference were all significantly decreased (p<0.0001) after the surgery.

**Table 1:**
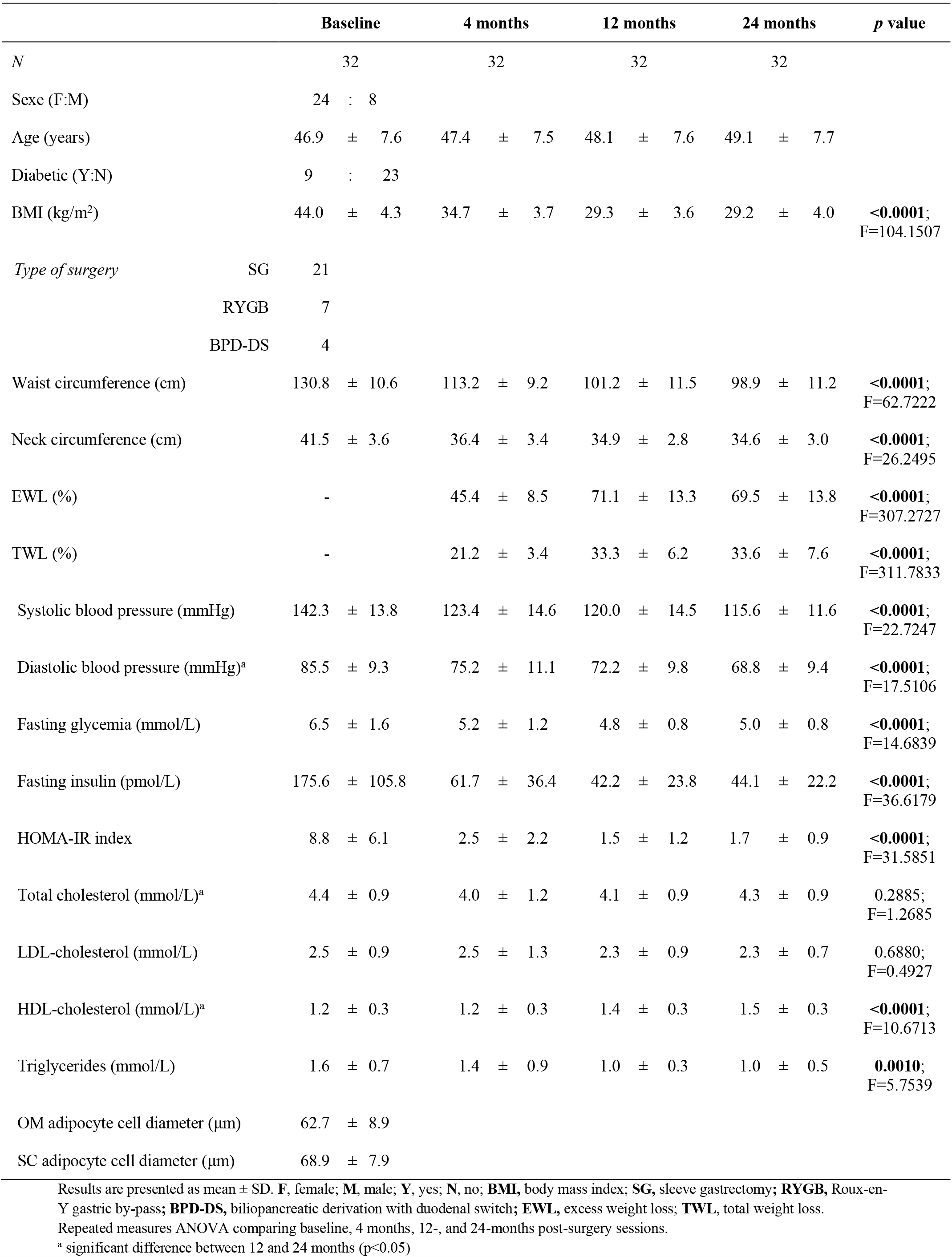
Characteristics of participants at baseline, 4, 12 and 24 months after bariatric surgery.

Most participants underwent sleeve gastrectomy. The mean total weight loss 24 months after surgery was 33.6 ± 7.6%. All metabolic markers significantly improved after the surgery [i.e. a decrease of systolic and diastolic blood pressure, fasting glycemia and fasting insulin, and HOMA-IR index, as well as most lipid profile markers (i.e. HDL-cholesterol and triglycerides)] as observed in **Table 1**. Only the total cholesterol and LDL-cholesterol did not improve significantly. Interestingly, no significant difference was observed in the mean BMI, total weight loss and the improvement in most metabolic markers between 12- and 24-months post-surgery (p>0.05). There was a significant difference for diastolic blood pressure (decrease at 24 months versus 12 months), total cholesterol and HDL-cholesterol values (increases at 24 months versus 12 months) (p<0.05).

### 3.2 Effect of bariatric surgery on WM and GM density

As shown in **Figure 1** and **Figure 2**, changes of voxel-wise WM and GM density previously observed at 12 months post-surgery [21] were still present at 24 months after the surgery. There was a widespread increase of WM density mainly in the cerebellum, the brainstem, and the corpus callosum (**Figure 1**). As for the GM density, there was also a widespread increase, but less extensive, mainly in the cerebellum, the occipital and temporal cortex, and the postcentral gyrus (**Figure 2**). No significant difference was found between surgery types or when comparing WM or GM densities between 12 and 24 months.

**Fig 1.**
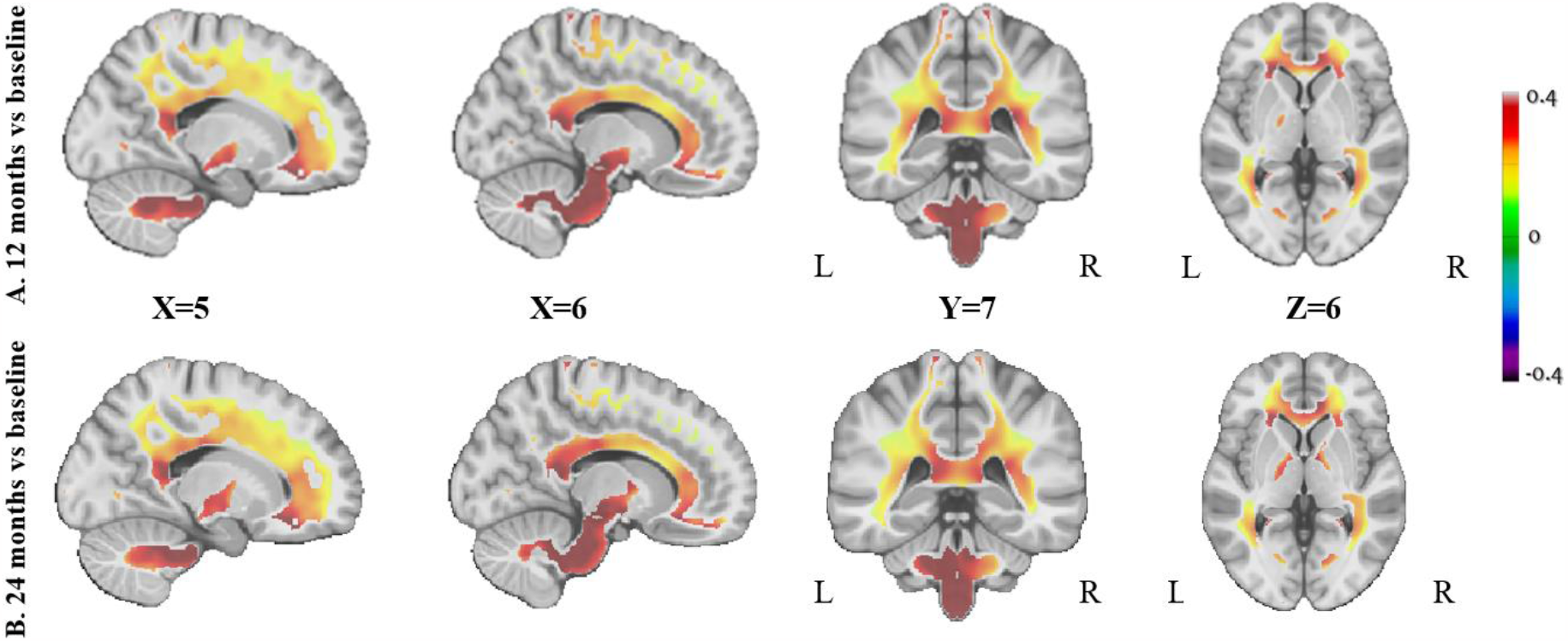
Changes in white matter (WM) density 12- and 24-months post-surgery compared to baseline. The figure shows the Beta value maps from the voxel-wise mixed-effects models for the WM regions that were significant after whole-brain FDR correction (p < 0.05), correcting for age at baseline, sex, initial BMI, and diabetic status. Colors show higher positive (in red) or negative (in dark purple) Beta values. L, left; R, right.

**Fig 2.**
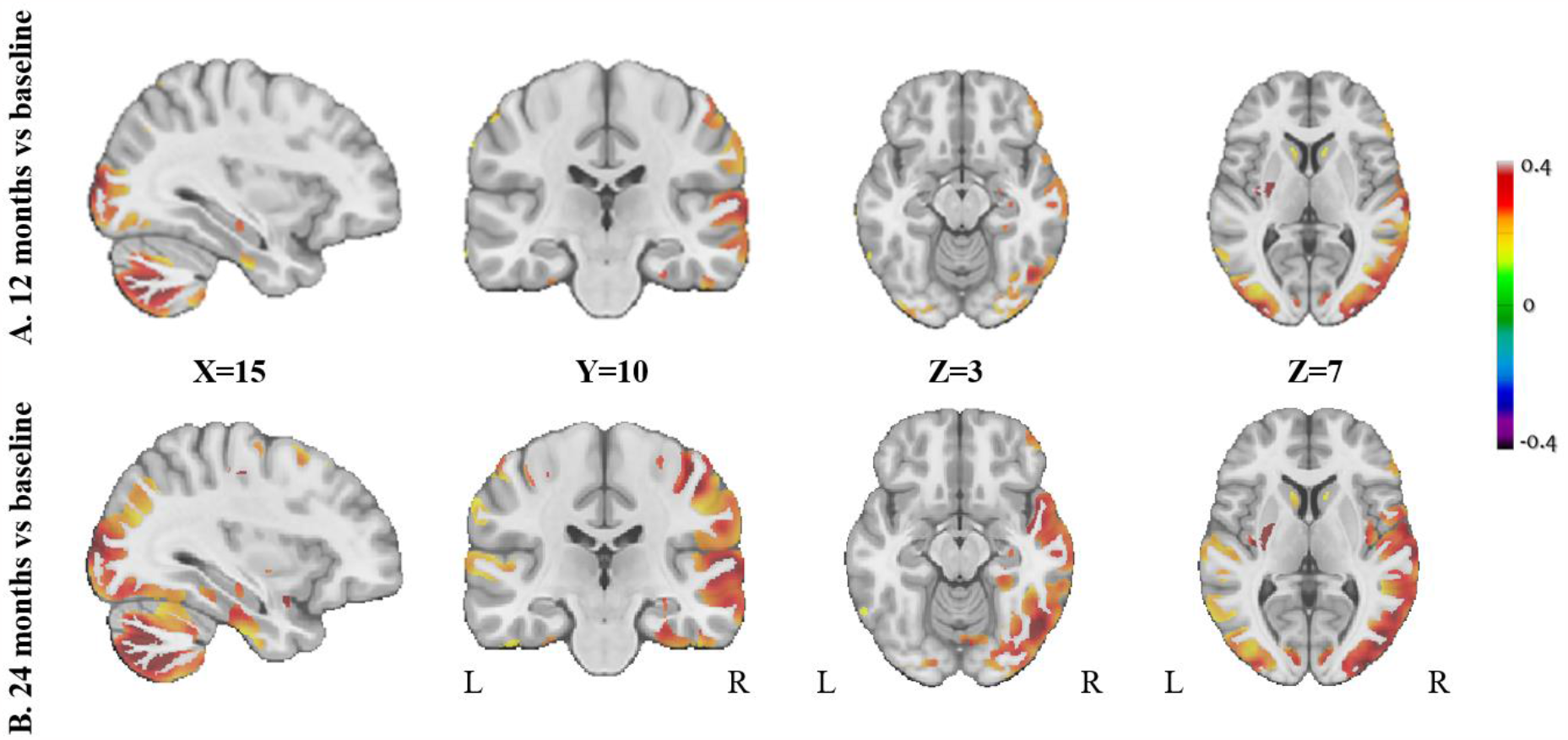
Changes in grey matter (GM) density 12- and 24-months post-surgery compared to baseline. The figure shows the Beta value maps from the voxel-wise mixed-effects models for the GM regions that were significant after whole brain FDR correction (p < 0.05), correcting for age, sex, initial BMI, and diabetic status. Colors show higher positive (in red) or negative (in dark purple) Beta values. L, left; R, right.

To make sure that the structural brain changes observed after bariatric surgery were not due to any confounders related to the repetition of the MRI session, a group of participants (n=19) had two pre-surgery sessions. The clinical characteristics of these participants were similar to our main cohort (**Table S1**). Participants were mainly women (74%) and the mean age at pre-surgery session 1 was 43.7 ± 10.4 years old. As expected, there was no significant difference in the BMI of these participants between the two pre-surgery sessions (p=0.9703, **Table S1**), and no significant changes were observed in the WM and GM densities.

### 3.3 Associations between changes in WM and GM densities and adiposity or metabolic markers following bariatric surgery

Linear mixed-effect models were used to assess the association between changes in WM or GM density and adiposity or metabolic measurements after bariatric surgery, accounting for age and sex. The adiposity markers used for the associations between changes in regional WM and GM densities were BMI, total weight loss, as well as waist and neck circumferences. Post-surgery reduction in adiposity measurements and total weight loss were significantly associated with increased WM density in 18 out of 27 regions, including acoustic radiation, central tegmental tract, cingulum, central nerve, corticospinal tract, corticostriatal and corticothalamic pathways, extreme capsule, fornix, frontopontine tract, inferior cerebellar peduncle, lateral lemniscus and pariotopontine tract (**Table S2**, p<0.0017). Regarding GM density, out of 34 regions, only increases in cingulate, rectus, precuneus, paracentral and vermis regions were significantly associated (**Table S3**, p<0.0014) with the reduction of one or more adiposity marker (BMI, total weight, waist and neck circumferences). A negative association was observed between total weight loss and increased GM density in occipital and angular regions (**Table S3**, p<0.0014).

**Tables S2** and **S3** show the associations between post-operative changes in metabolic markers (systolic blood pressure, levels of triglyceride and insulin as well as HOMA-IR index) and regional WM or GM densities, respectively. Significant associations were observed between post-surgery improvement in plasma insulin levels and HOMA-IR index and increased WM density in 4 regions, including the cingulum, fornix, inferior cerebellar peduncle and posterior commissure (p<0.0017 for all, **Table S2**). A few significant associations were observed between post-surgery reduction in systolic blood pressure or triglycerides levels and increased WM density in cingulum and inferior cerebellar peduncle (p≤0.0017 for all). Increased GM density in occipital and angular regions was significantly associated with higher post-surgery systolic blood pressure (p<0.0014, **Table S3**), while significant associations were observed between post-surgery improvement in plasma insulin levels and HOMA-IR index and increased GM density in rectus and superior motor area regions (p<0.0014, **Table S3**).

### 3.4 Associations between abdominal adipocyte diameter and changes in WM and GM densities following bariatric surgery

Mean adipocyte diameter of the 29 participants from which tissue was collected at the time of surgery was 62.7±8.9 μm for the OM adipose tissue, and 68.9±7.9 μm for the SC adipose tissue. Spearman correlations were performed to examine the association between abdominal OM or SC adipocyte diameter and adiposity or metabolic markers. A positive and significant association was found between OM adipocyte diameter and neck circumference at baseline (0.5141, p<0.0001). A positive and significant association was found between OM adipocyte diameter and insulin levels (0.3809, p=0.0017), while a positive and significant association was observed between SC adipocyte diameter and triglyceride levels (0.3920, p=0.0012) as well as HOMA-IR index (0.3201, p=0.0093). No significant association was observed between OM or SC adipocyte diameter and the percentage of total weight loss at 24 months (p=0.4184 for OM; p=0.9666 for SC).

We used linear mixed effect models to examine if adipocyte diameter (OM or SC) at the time of surgery was associated with post-surgery global GM and WM density changes, accounting for age and sex. The variable of interest was an interaction term between session and OM or SC, indicating whether those with higher baseline OM or SC values experienced greater change in the VBM measurements at each session. We found a significant positive association between pre-operative OM adipocyte diameter and changes in total WM density at 24 months (t=2.6895, p=0.008), suggesting that higher OM adipocyte diameter was associated with greater changes in WM density after 24 months. A marginal positive association was found between OM adipocyte diameter and total WM density after 4 (t=1.8065, p=0.07) and 12 months (t=1.4476, p=0.15). A trend was observed between pre-operative SC adipocyte diameter and changes in total WM density at 24 months (t=1.9748, p=0.05). A marginal positive association was found between SC adipocyte diameter and total WM density at 4 months (t=1.5611, p=0.12). However, no significant association was observed between SC adipocyte diameter and total WM density at 12 months (t=0.5517, p=0.58). No significant association was found between OM or SC adipocyte diameter and total GM density at 4, 12 or 24 months. The results remained similar when including diabetic status, baseline BMI, and surgery type as covariates in the models.

## 4. Discussion

In this study, we demonstrated that the increases in WM and GM densities previously observed 12 months post-surgery [21] are still present up to 24 months post-surgery and in the same brain regions. As previously reported, these increases were more significant and extended for WM compared to GM. The increased densities, mainly in WM regions, were significantly associated with the magnitude of weight loss and metabolic improvements. We also found that larger OM adipocyte diameter at the time of surgery, which is known as a marker of adipose tissue dysfunction, is associated with greater increases in WM density 24 months after surgery. Furthermore, to verify that the changes in WM and GM densities observed after bariatric surgery were not due to any confounds related to the repetition of the MRI session and were related to weight-loss induced-bariatric surgery, a separate group of participants underwent two scanning sessions before the surgery. As expected, no significant changes in GM and WM densities were observed in the two MRI sessions performed before surgery, suggesting that the neuroanatomical increases observed after the surgery are related to the weight loss and cardiometabolic improvements induced by the surgery.

To the best of our knowledge, we are the first to report that the increases in WM and GM densities observed after bariatric surgery persist at 24 months. More specifically, we found widespread increases in WM density, particularly in cerebellum, brainstem, and corpus callosum, as well as less extended increases in GM density, particularly in the cerebellum, the occipital and temporal cortex, and the postcentral gyrus. These results are in line with previous studies from our group [21] and others [23]– [25] with shorter follow-up (less than 12 months). Interestingly, we previously found that these post-operative structural brain changes were spatially overlapped with brain differences between individuals with severe obesity and those with normal body mass index (where those with obesity had significantly lower VBM densities than matched controls), supporting the hypothesis that the effects of bariatric surgery represent a resolution of adiposity-related brain alteration, even after 24 months [21].

The associations we found between the post-surgery increase in WM density and the improvement of metabolic and adiposity markers are also consistent with the literature and our previous studies. Indeed, the post-surgery increases of WM density as well as resting neural activity (as measured by fractional-amplitude of low frequency fluctuations) were significantly associated with the amount of weight loss and improvement of the metabolic/inflammatory profiles [21]–[25]. Furthermore, the increase of WM and GM densities were observed in the same regions 24 months and 12 months post-surgery, and there was no significant difference in the WM and GM densities between the two sessions. This could be explained by the fact that the percentage of weight loss and metabolic improvements are similar between these two sessions. The widespread increases in GM density are also in line with our recent study [26] showing a significant improvement in brain health following bariatric surgery, as indicated by an impressive decrease of 2.9 and 5.6 years in brain age 12 and 24 months post-surgery, respectively. Taken together, this improvement in brain integrity and brain health following bariatric surgery could be a consequence of metabolic and vascular improvement induced by weight-loss surgery.

The underlying mechanisms explaining the increase in WM and GM densities after bariatric surgery remain unknown. One of the potential mechanisms to explain these results is the improvement of the chronic, low grade inflammatory state observed after bariatric surgery-induced weight loss, which can affect brain vasculature and blood-brain barrier permeability [38]–[40]. Indeed, it is possible that the structural changes identified with MRI derive from non-neuronal components of the brain, such as the vasculature, which accounts for about 5% of GM [38]. Interestingly, a recent study found that bariatric surgery is associated with reduced brain glucose utilization, which is directly related to improvements in cognitive performance [41]. These improvements of glucose and insulin homeostasis, reduction of inflammation and improvements in cerebral blood flow observed after bariatric surgery [42]–[44] may lead to angiogenesis, neurogenesis, gliogenesis, axon sprouting and synaptic remodeling, which could influence WM and GM densities [38], [45]. An increase of WM density could also be caused by a higher number of myelinated axons in a tract or a higher thickness of myelin [38], although such changes would seem unlikely to happen over the time periods studied. Future studies are needed to better understand the mechanisms underlying these structural changes and their link with cognitive performance.

We also report that individuals with higher OM adipocyte diameter at the time of surgery had greater changes in total WM density 24 months post-surgery. Similar associations were observed with SC adipocyte diameter, but to a lesser extent. However, the abdominal adipocyte diameters were not related to the changes in GM density. This is consistent with the fact that the associations between the improvement of adiposity/metabolic markers and post-surgery increase in brain densities were mainly in WM. A recent review by Garcia-Garcia [17] reported that obesity-induced inflammation is related to disruptions in WM integrity and cerebrovascular disease. Such systemic, low-grade inflammation might be the result of adipose tissue dysfunction which is characterized by adipocytes hypertrophy [5]. Indeed, in response to sustained positive energy imbalance, adipose tissue may reach a level at which adipocytes become dysfunctional, which limits further lipid storage in adipose tissues and leads to ectopic fat deposition and cardiometabolic alterations. Typically, hypertrophy is more common within OM adipose tissue than SC adipose tissue, where the expansion of adipose tissue takes place mostly through the generation of new adipocytes (adipose tissue hyperplasia) [2], [3]. When adipocytes are hypertrophic, an elevated secretion of proinflammatory cytokines is observed, which can have detrimental effects on multiple organs, including the brain [17], [46]. Previous studies have found that this inflammation was reduced following bariatric surgery among many other metabolic variables [19]. It is also important to note that WM could be more sensitive to inflammation changes observed after bariatric surgery [23], possibly explaining why we only observed associations between adipocyte diameter and WM density, and not GM density. Because participants with larger OM adipocyte diameter in our study had greater increases in total WM density 24 months post-surgery, this suggests that individuals with abdominal obesity might benefit the most from the bariatric surgery at the neural level.

Some limitations should be acknowledged. First, we did not include a control group of individuals with normal weight with 24 months of follow-up to assess if the changes that we found at 24 months follow-up would be observed in a population without obesity. However, we added a separate group of participants with severe obesity who underwent two scanning sessions before the surgery to make sure the effects seen after bariatric surgery were not due to any confounds or artifactual T1 signal changes related to the repetition of the MRI session. Another limitation is that we did not have adipose tissue samples for 3 participants. Even if we included the surgery type as a covariate in our statistical models, it was not possible to compare the different surgical interventions because our sample sizes were too small for two surgical groups (biliopancreatic diversion with duodenal switch and Roux-en-Y gastric bypass). Another limitation is that we used VBM to assess WM density, but diffusion-weighted imaging would have been a more sensitive method for evaluating changes in WM integrity [47].

In conclusion, our results show extended increases of WM density up to 24 months post-surgery which are related to metabolic and adiposity improvements. Greater pre-operative OM adipocyte diameter is associated with greater increase in WM density at 24 months, suggesting that individuals with higher abdominal adiposity might benefit the most from surgery. More research is needed to understand the underlying mechanisms explaining the brain structural changes after weight loss induced by bariatric surgery.

## Data Availability

All data produced in the present study are available upon reasonable request to the authors.

## Author contributions

Conceptualization: LB; SF; AT; AD; DR; MD; AM

Methodology: AM, MD, ML, AT, MFG

Data collection (participants): MP, ML, LB

Data analysis: ML, MFG, AL, MEL, YZ, MD

Writing – original draft preparation: ML, MEL, MFG, AM

Writing – review and editing: SI; YZ; LB; SF; AT; AD; DR; MD; AM

Supervision: AM, MD

Funding: AT, LB, DR

