## Supplemental Table 1 for "Sustained improvements in brain health and metabolic markers 24 months following bariatric surgery"

### Supplemental material

#### Table S1: Characteristics of the group of participants who had two pre-surgery sessions

| | Baseline 1 | Baseline 2 | *p* value |
|---|---|---|---|
| *N* | 19 | 19 | |
| Sexe (F:M) | 13 : 6 | 13 : 6 | |
| Age (years) | 43.8 ± 10.2 | 43.9 ± 9.0 | 0.9884 ; t=2.02619 |
| Diabetic (Y:N) | 6 13 | 6 : 13 | |
| BMI ($kg/m^2$) | 41.7 ± 3.8 | 42.0 ± 3.6 | 0.9703; t=2.02809 |
| Waist circumference (cm) | 126.2 ± 10.5 | 127.1 ± 11.0 | 0.8000 ; t=2.02619 |
| Neck circumference (cm) | 41.7 ± 3.8 | 41.8 ± 4.3 | 0.9286 ; t=2.02619 |
| EWL (%) | - | -0.008 ± 0.04 | 0.3037; t=2.02809 |
| TWL (%) | - | -0.001 ± 0.02 | 0.4885 ; t=2.02619 |
| Systolic blood pressure (mmHg) | 128.0 ± 12.7 | 123.5 ± 12.6 | 0.2844 ; t=2.03011 |
| Diastolic blood pressure (mmHg) | 76.2 ± 11.9 | 74.0 ± 10.2 | 0.5551 ; t=2.03011 |

Results are presented as mean ± SD. **F**, female; **M**, male; **Y**, yes; **N**, no; **BMI,** body mass index; **SG,** sleeve gastrectomy**; RYGB,** Roux-en-Y gastric by-pass**; BPD-DS,** biliopancreatic derivation with duodenal switch**; EWL,** excess weight loss; **TWL**, total weight loss.
